# Estimation of global case fatality rate of coronavirus disease 2019 (COVID-19) using meta-analyses: Comparison between calendar date and days since the outbreak of the first confirmed case

**DOI:** 10.1101/2020.06.11.20128959

**Authors:** Ramy Abou Ghayda, Keum Hwa Lee, Young Joo Han, Seohyun Ryu, Sung Hwi Hong, Sojung Yoon, Gwang Hun Jeong, Jinhee Lee, Jun Young Lee, Jae Won Yang, Maria Effenberger, Michael Eisenhut, Andreas Kronbichler, Marco Solmi, Han Li, Louis Jacob, Ai Koyanagi, Joaquim Radua, Jae Il Shin, Lee Smith

## Abstract

Since the outbreak of the coronavirus disease 2019 (COVID-19) in December of 2019 in China, the estimation of the pandemic’s case fatality rate (CFR) has been the focus and interest of many stakeholders. In this manuscript, we prove that the method of using the cumulative CFR is static and does not reflect the trend according to the daily change per unit of time. A proportion meta-analysis was carried out on CFR in every country reporting COVID-19 cases. Based on the results, we performed a meta-analysis for global COVID-19 CFR. Each analysis was performed on two different calculations of CFR: according to calendar date and according to days since the outbreak of the first confirmed case. We thus explored an innovative and original calculation of CFR concurrently based on the date of the first confirmed case as well as on a daily basis. For the first time, we showed that using meta-analyses, according to calendar date and days since the outbreak of the first confirmed case were different. We propose that CFR according to days since the outbreak of the first confirmed case might be a better predictor of the current CFR of COVID-19 and its kinetics.

## 1. Introduction

Since the outbreak of the coronavirus disease 2019 (COVID-19) in December of 2019 in China, COVID-19 has spread worldwide [1,2]. As of May 28^th^, 2020, 5,593,631 confirmed cases with 353,334 deaths were reported across the 216 affected countries, territories, or areas [3]. Among other clinical and epidemiologic features of the virus, predicting the estimates of mortality of this pandemic is crucial and indispensable.

The estimate of the case fatality rate (CFR), is defined as the number of deaths with COVID-19 divided by the number of confirmed COVID-19 cases. CFR has been developed to understand the mortality and epidemiological features reporting for emerging infectious diseases [4,5], such as Severe Acute Respiratory Syndrome-coronavirus (SARS, CFR 9.6% on a global scale) [6] and Middle East Respiratory Syndrome-coronavirus (MERS, CFR 34.5%) [7]. To date, there have been many attempts to estimate the underlying “true CFR” of COVID-19 [8-13]. However, these CFR are not without limitations. These estimates need to be treated with extreme caution because each region of the world is experiencing a different stage of the pandemic. In addition, CFR is contingent on many other factors, including the extensiveness detection and testing efficiency, local health and pandemic response policies, and the condition and inclusiveness of the already existing health systems. Failure to consider these former factors and simply dividing the cumulative deaths with COVID-19 by cumulative confirmed cases based on the latest global statistics available will inevitably distort the CFR in each stage of COVID-19 into an unknown direction, let alone fail to reveal the true dynamics of CFR of the disease. In addition, several previously published papers [14,15] suggested models using CFR should be based on the cumulative confirmed cases and deaths with a simple linear regression analysis. However, this method of using the cumulative number is static and does not reflect the trend according to the daily change per unit of time. Additionally, it prevents an exact estimation of CFR because the number of the confirmed cases and the onset time of the first case vary by country, and even within regions of the same country.

Therefore, to get as close as possible to a real estimate, we calculated the CFR of each country, concurrently based on the date of the first confirmed case as well as on a daily basis.

## 2. Materials & Methods

Proportion meta-analyses were performed to obtain the average CFR for each day, commencing from the date of the first confirmed case to the present, stratified by each country. Therefore, we present unique CFR dynamics obtained by correcting and theoretically circumventing the bias created by the fact that each country is facing different stages of the pandemic. This approach to the CFR provides a new insight that lays the foundation for a proper analysis of CFR. One caveat that we acknowledge is that many potential positive cases that were not tested might present possible confounding variables, skewing our results in a specific direction. At this point, it is impossible to account for the totality of the COVID-19 cases (tested and not tested), and this calculation is out of the scope of this study.

Global data of COVID-19 confirmed cases and deaths with COVID-19 were collected from the European Centre for Disease Prevention and Control (ECDC, https://www.ecdc.europa.eu/en/geographical-distribution-2019-ncov-cases), which showed each country’s data from Dec 31, 2019, to May 8, 2020. The CFR was defined as follows:

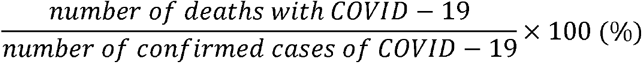

To note, our analysis of CFR included only confirmed cases by molecular or serological testing. The data had blanks since the reports from each country were not continuous on a daily basis, especially during the early stages of the epidemic secondary to under-testing and under-reporting of cases. After multiple rounds of discussions, for calculation simplicity, the blanks were decided to be processed as the number of cases in the most recent report before the blank rather than splitting the number of cases equally among the missing days. We stratified the confirmed and deaths of COVID-19 for each country according to days since this country reported its first confirmed case of COVID-19.

A proportion meta-analysis was then carried out on CFR in every country reporting COVID-19 cases. Based on the results, we performed a meta-analysis for global COVID-19 CFR. Each analysis was performed on two different calculations of CFR: according to cases calendar date and according to days since the outbreak of the first confirmed case. Every analysis was based on reports until May 8, 2020.

For a meta-analysis of the CFR of COVID-19, MedCalc version 19.2.1 software (MedCalc Software, Ostend, Belgium, trial version) was used to analyze the summary effects with 95% confidence interval (CI) and between-study heterogeneity. We performed a proportion meta-analysis to estimate the summary effects. The summary effects obtained by the proportion meta-analysis of the CFR under the fixed- and random-effect model for each data over time were presented as figures, and the 95% Confidence Interval, CI, are summarized in the Supplementary Table 1 and 2. To determine the extent of variation between the studies, we did heterogeneity tests with Higgins’ I^2^ statistic^14^. An I^2^ value below 50% represented low or moderate heterogeneity, while I^2^ >50% represented high heterogeneity [16]. For graphing the patterns of CFR in all countries, Microsoft Excel version 2013 was used.

## 3. Results

### 3.1 The new dynamics of CFR revealed after the meta-analyses

Figure 1 presents the following data over time: the fixed- and the random-model results of the meta-analysis, the pooled estimate, and the number of total cases included in each analysis.

**Figure 1.**
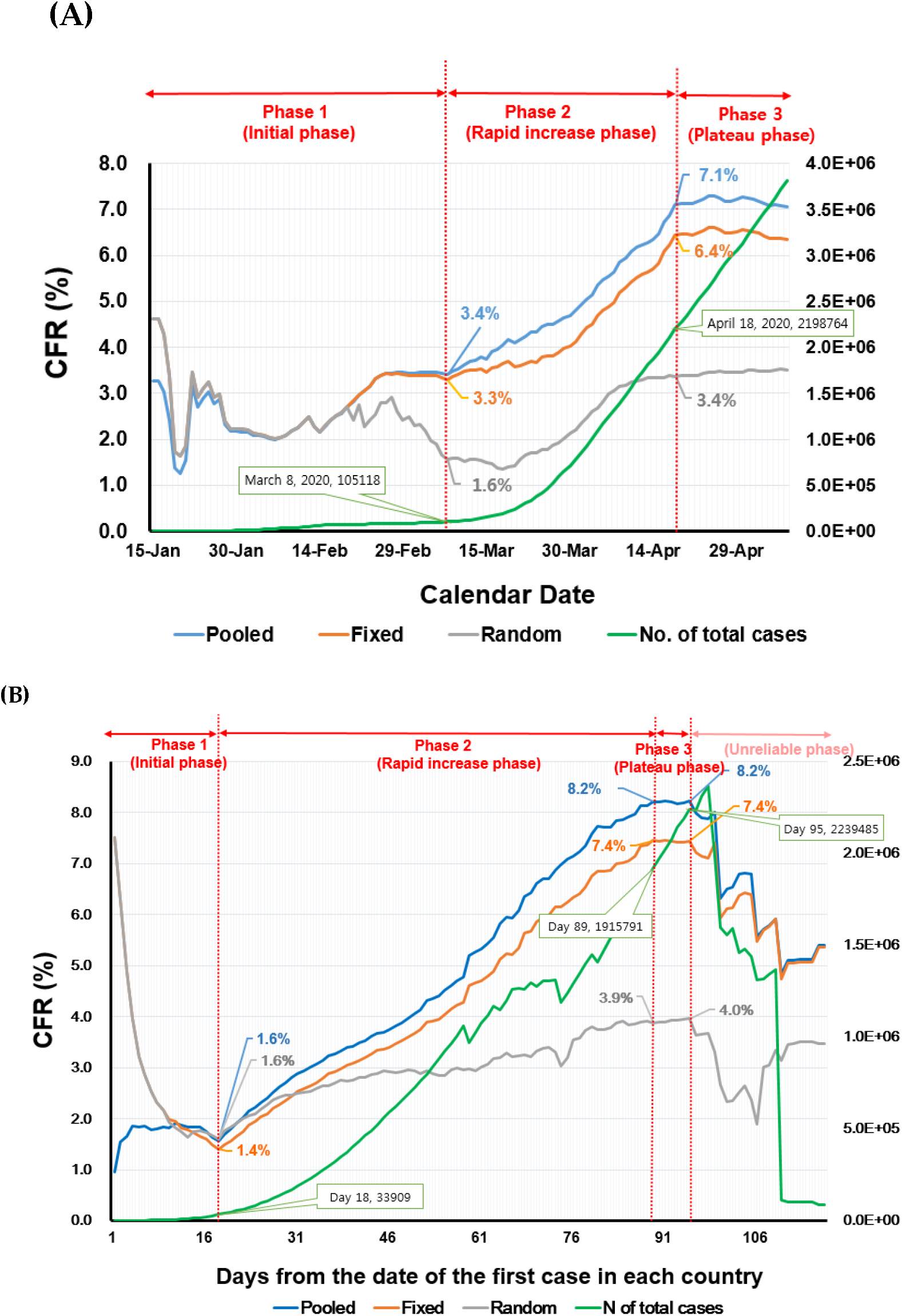
Timeline of CFR in worldwide among countries with COVID-19 reports until Apr 17, 2020: (A) According to date and (B) According to days since 1^st^ confirmed case. COVID-19: Coronavirus 2019, CFR: case fatality rate, fixed: fixed-effect model, random: random-effect model, pooled: calculated CFR based on incidence and mortality data, N: number

Figure 2 presents the trend of patients with COVID-19 according to date and according to days. We obtained the time trend of CFR by calculating pooled estimates, fixed- and random-effect estimates from meta-analyses by the calendar date and by days since the first confirmed case

**Figure 2.**
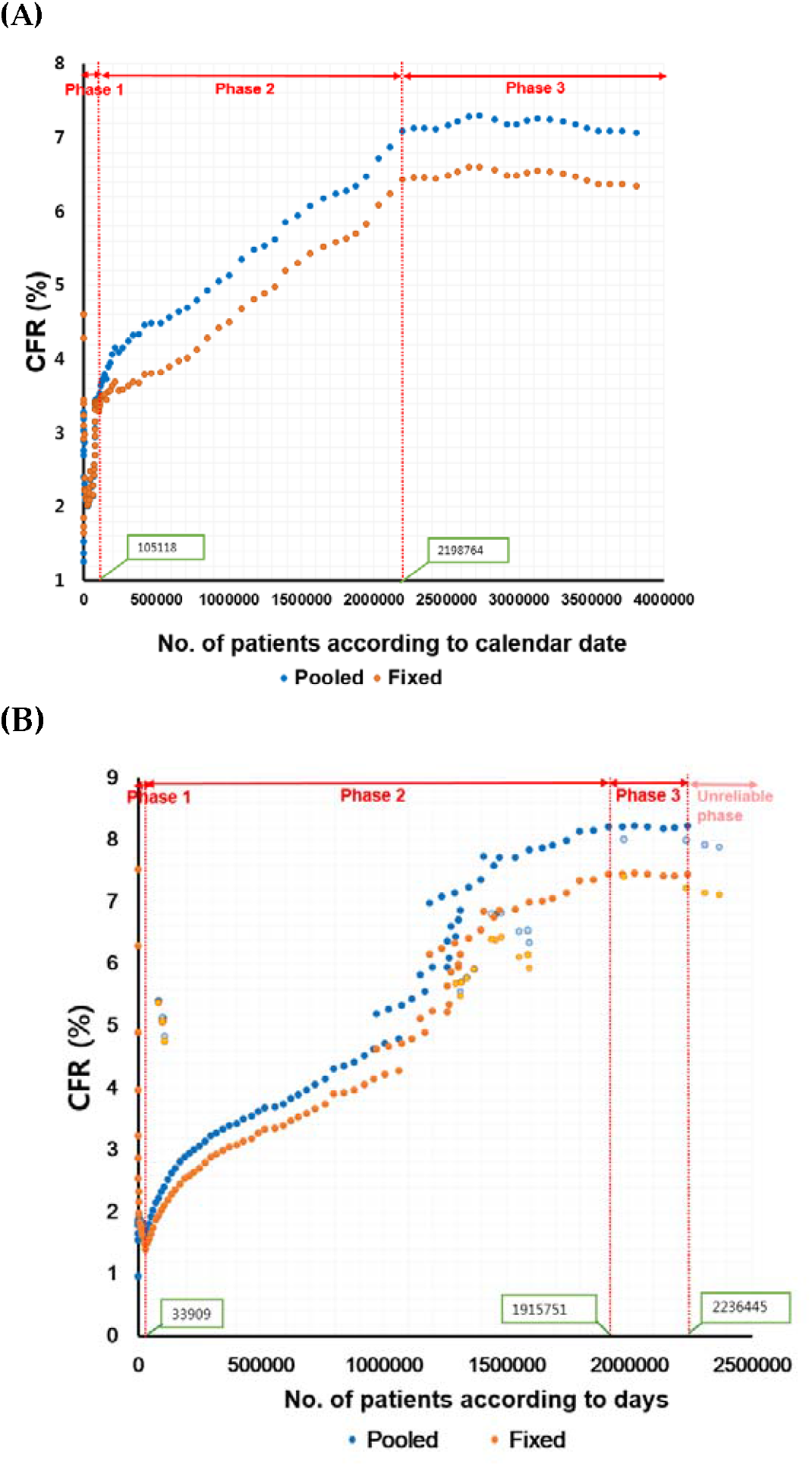
The trend of patients with COVID-19: (A) According to date and (B) According to days. COVID-19: Coronavirus 2019, CFR: case fatality rate, No.: number, fixed: fixed-effect model, pooled: calculated CFR based on incidence and mortality data

By comparing the figures that show the time trend of CFR stratified by the two methods, it was visually observed that the CFRs calculated by each sorting method had different trends over time: Figure 1A (CFR stratified by calendar date) vs. Figure 1B (CFR stratified by days since the first confirmed case). In both figures, results from the random- and the fixed-effect model were almost identical; however, after they diverge, the fixed-effect model was similar to the pooled estimates while the random-effect model estimates were smaller. One possible explanation for the fact that random CFR estimates were lower than the fixed estimate is that less weight is given to countries with a small number of confirmed cases compared to countries with a high number of cases. Namely, a higher weight is given to countries with a large number of confirmed cases in the random estimate. On the other hand, lower weight is given to countries with a low number of cases in the fixed estimate (Figure 3A and 3B).

**Figure 3.**
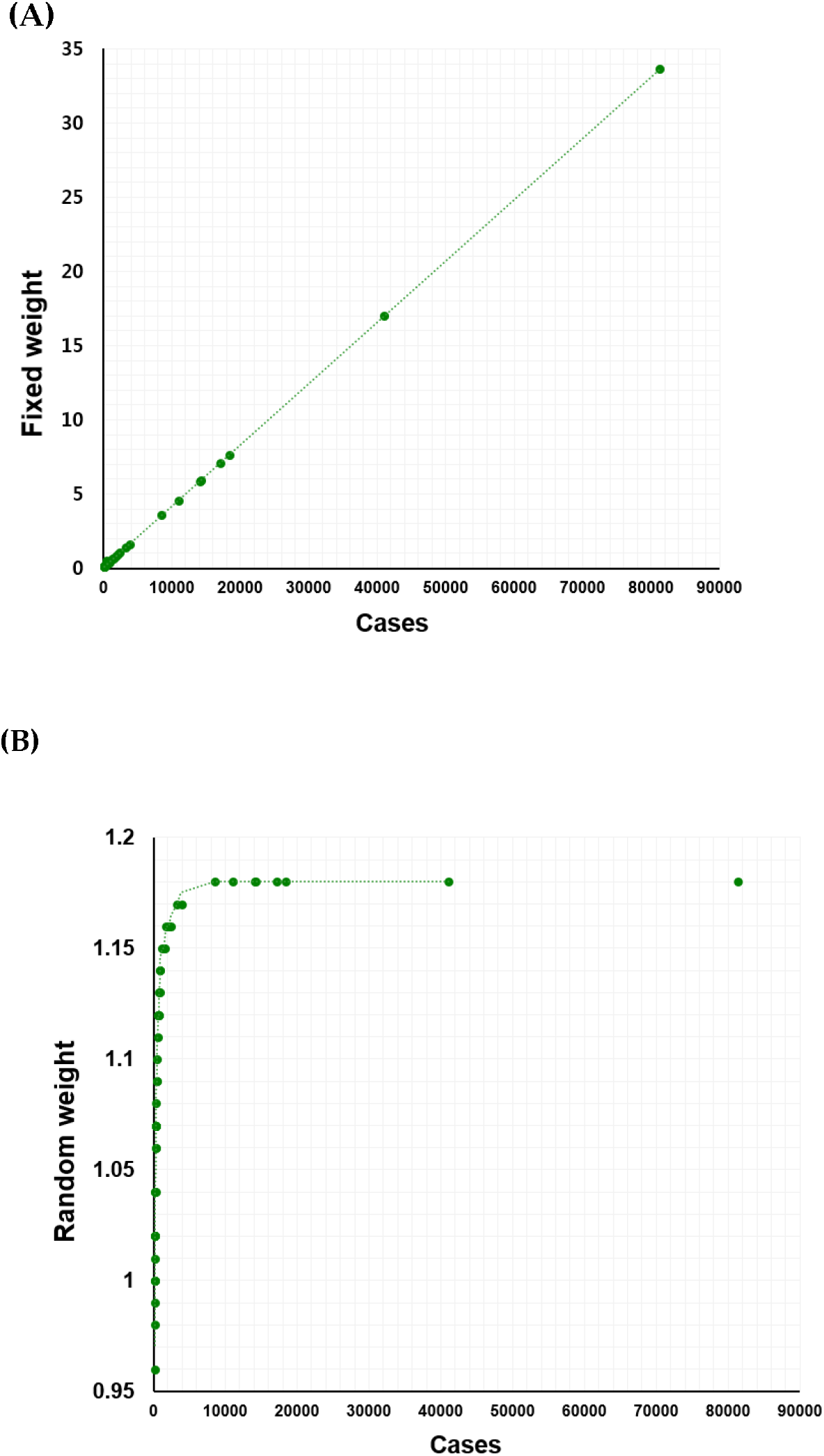
Differences in weight between (A) Random and (B) Fixed meta-analyses to the number of patients at one point as an example (50 countries, March 20, 2020)

We identified 4 distinct phases based on our results. In Figure 1A, phase 1 contains data from January 15 to March 7, 2020, phase 2 included data from March 8 to April 17 and phase 3 was from April 18 to May 8. In phase 1, all CFRs ranged between 1% and 3%. However, from March 8 to April 18 (phase 2), both fixed and pooled CFRs increased rapidly from 3.29% to 6.43% for fixed-effect CFR and 3.40% to 7.09% for pooled CFR. From April 18th with 2,198,764 confirmed patients to May 8th, both fixed and pooled CFRs remained at 6%p and 7%, respectively (phase 3). Interestingly, in phase 3, the CFR remains similar even though the number of confirmed patients per date continues to rise after a total of 2,198,764 was reached. Figure 2B shows a similar trend with the characteristic’s phases 1 to 3, parallel to those in Figure 1. From the day the first COVID-19 case was confirmed or Day 1 to Day 17, the CFR ranged between 1.4 and 1.6% (phase 1). It showed an exploding increase from Day 18 to Day 88, when the number of confirmed patients per day reached 33,909 (phase 2). As the number of confirmed patients increased to 1,915,791 on Day 89, the fixed CFR remained at 7.45% and the pooled CFR remains at 8.21%, despite the fact that the number of confirmed patients increased rapidly. Importantly, in Figure 2B, an “unreliable phase” was added, which occurred after Day 95, because the number of the confirmed cases and deaths included in the analysis decreased significantly, due to the long-lived epidemic in only a few countries such as China, Italy or Spain.

Comparing Figure 1A and 1B, we found that both pooled and fixed CFRs increased approximately 1%P after adjusting the CFR standard to the days since the first confirmed case (7.09% to 8.20%, 6.40% to 7.40%, respectively). Therefore, the CFR in the plateau phase was approximately 1%P higher in the meta-analyses by days since the first confirmed patient compared to the meta-analyses by date. This might be explained by the “noise” in the data from the early days of the epidemic in each country. Analogous comparison of Figure 2A and 2B revealed a similar 1%P approximate increase in phase 3, the plateau phase, between CFR by days since the first confirmed patient compared to the meta-analyses by date.

An additional phase emerged in Figure 1B since countries with newly emerging COVID-19 moved to the front of onset period as they were sorted by days (Supplementary Figure 2, see Appendix). Similarly, I^2^ value earlier reached above 50% representing high heterogeneity was at Day 15 and showed a plateau pattern since Day 50 according to days (Supplementary Figure 1B) than according to calendar date reached above 50% on February 25, 2020 and marked plateau since April 12, 2020 (Supplementary Figure 1A, see Appendix).

Figure 1B identifies the time period of the day 15 in which the fixed- and random-model estimates split and the estimates of fixed-model proceeds in a similar direction to the pooled model estimates.

### 3.2 Correlations between the number of confirmed patients and CFR

Based on Figure 1, we also investigated the relationship between CFR and the number of confirmed COVID-19 patients (Figure 2). Figure 2A was devised using the number of patients according to the calendar date rather than the cumulative number of patients. This figure revealed that CFRs linearly correlated with the number of confirmed cases, the more the number of confirmed cases, the higher the CFR. On the other hand, when the number of patients was adjusted by days since the first confirmed case, as shown in Figure 2B, CFR increases, as shown in Figure 1, until the number of confirmed patients per day reaches 1.0 million cases. Following this phase, CFR then rapidly increases between 1.0 million and 1.5 million cases. After 2.0 million cases, a plateau pattern continues.

In Figure 2B, the blurry dots represent CFRs in the “unreliable phase.” In this phase, CFR decreases when the number of confirmed patients falls below 2 million. The unreliable phase could represent potentially a new phase, the decreasing phase. The model according to calendar date (Figure 2A) may have underestimated the CFR, this might be because countries being in different stages, and thus phases, of the disease.

## 4. Discussion

We propose that CFR is not a fixed, static rate. It is rather dynamic, constantly fluctuating with time, location, and population, as confirmed in Figure 1. In this context, it is important to view CFR as a function of time, rather than presenting CFR as a single and absolute value. Stratifying CFR by days since the first confirmed case is a novel and innovative attempt to uncover the dynamics of CFR as the epidemic unfolds itself. We believe that the CFR simply stratified by calendar date does not reflect the true epidemic situation of each country.

Our analysis revealed a CFR trend consisting of four distinctive phases. Based on our results, we carefully propose that the slope of epidemic model will proceed to the next four stages as follows: phase 1 or initial phase, phase 2 or rapid increase phase, phase 3 or plateau phase and phase 4 or decreasing phase. Based on this statistical trend, it is estimated that the global situation of the pandemic will slow down from the time all countries reach phase 3, and it can be improved when the situation has reached the end of the phase. However, as mentioned above and analyzing the data of 100 days so far, the world may remain in phase 3 as of May 2020 for an undetermined amount of time, and CFR may not have yet reached phase 4. It may take considerable amount of time to enter this final phase.

The method of calculating CFR needs to be cautioned, and its limitations acknowledged. The numerator and the denominator of CFR should be composed of patients infected at the same time as those who died to accurately represent the CFR. To overcome this restraint, Baud et al.[8] and Wilson et al.[9] proposed time delay-adjusted CFR to correct the delay between confirmation and death. They adjusted the denominator of CFR as the number of confirmed cases 13-14 days before the measured date to calculate the number of confirmed cases infected concurrently to those who died. Based on these articles, researchers at Oxford University used their global COVID-19 CFR model according to the date since the outbreak in Jan 2020 [17]. However, Oxford’s calculation is also flawed since 13 to 14 days before the date of test confirmation is not necessarily the date when a subject is infected [12]. Moreover, there are cases that show test positivity even after recovery. Additionally, the stretching and overwhelming of the healthcare systems creates a delay between testing and receiving the results, thus confirming the case. As this adjusted time-delay CFR leads the estimate to an unknown bias [12, 13], we used the conventional method to calculate CFR. Moreover, the numerator of the CFR is the number of deaths with COVID-19. We should be aware that this number is imperfect, and may include deaths not directly caused by COVID-19, such as fatal comorbid diseases. This may lead to an overestimation of the number relative to its true value.

In the present study, we observed unusually exaggerated estimates from our meta-analyses in the early phase (Phase 1) of the epidemic, both in CFR based on the calendar date and days since the first confirmed case. This is thought to be a statistical bias, as many groups and countries with small numbers were included. The studies included in the early phase of the epidemic are mostly a bundle of data in which deaths sporadically occurred in very small group sizes. Such data distribution may have severely exaggerated the meta-analyses results. Therefore, we believe that we should aim for a more standardized and homogenous analysis of the numbers. One method would be to observe the results from the time when the number of confirmed cases in each country has reached a certain distinct level. As a criterion for the time when the result of the meta-analyses becomes meaningful (the starting period of Phase 2), we presented the time when the fixed-effect model and random-effect model coincided: in the time-trend graph of every meta-analysis, the initial estimates of the random-model and the fixed-model almost coincides, and diverges from a certain point of time which is interestingly Day 15 from the first case in each country.

There are inevitable errors that arise because actual confirmed cases or COVID-19 deaths are not properly reflected due to differences in the medical system capabilities and the response to the pandemic in each country. Moreover, the number of screening tests for COVID-19 differs by the diverse screening criteria of each country. The rapid spread of COVID-19 means there are cases and deaths that not accounted for and consequently not recorded in the reported statistics. The screening criteria may have changed and evolve as COVID-19 spreads in a country. For example, South Korea had originally limited the screening tests to people with fever (37·5°C or above), respiratory symptoms who had contacts with a person returning from China or confirmed symptomatic cases within the last 14 days [18]. But as the disease spreads, South Korea widened the screening criteria to anyone who needs to be hospitalized based on a physician’s decision due to pneumonia from an unknown source [18].

The capability and efficiency of healthcare systems and the testing ability of COVID-19 are also important. When the overall testing capability for COVID-19 is limited to the most severe cases hospitalized with severe illness, the number of confirmed COVID-19 cases with mild symptoms and easily recoverable cases will be underrepresented resulting in artificially inflated CFRs. The capability of healthcare systems is central because, in practice, there are many cases where COVID-19 patients are concentrated in one geographical location. Cases of the disease in Italy spread fast, especially in the North, with an overwhelming proportion of individuals in need of intensive care units [19,20].

Through our meta-analysis of CFR calculated from the first confirmed case, we set new standards for observing CFR and suggest the four phases of epidemic pattern. From the results, it is unclear whether the CFR time trend could be explained by our epidemic stages of COVID-19. This might be because the pandemic is still in progress and the data is far from complete and accurate. Future studies and discussions are needed to fulfill the unmet need for a consensus of the definition of each phase. It would also be interesting to explore the relation between CFR and the number of testing performed. More specifically, it would be of great added value to explore if higher number of testing and availability is associated with a lower CFR. When the CFR is estimated by day since the first confirmed case, the estimates could be more representative of “the true kinetics” of COVID-19 CFR by time in a country. The stages of the epidemic should be classified by setting appropriate standards based on reliable global data, and discussion of setting these standards is necessary. It is noteworthy to say that since the COVID-19 is an ongoing and unfolding pandemic, we caution that the CFR time trend according to calendar days since the first confirmed case cannot be used to predict future CFRs.

## 5. Conclusion

This report highlights that the CFR is not a fixed value, rather it is dynamic, and it changes with time and population. Therefore, we strongly urge caution when dealing with CFR values, especially in an ongoing epidemic. We originally showed that estimation of global CFR of COVID-19, using meta-analyses of CFR, according to the calendar date and days since the outbreak of the first confirmed case were different. We propose that CFR according to days since the outbreak of the first confirmed case might be a better predictor of the current CFR of COVID-19 and its kinetics.

## Data Availability

All data is available on the WHO and European Centre for Disease Prevention and Control (ECDC) https://www.ecdc.europa.eu/en/geographical-distribution-2019-ncov-cases

## Author contribution

Conceptualization: JIS, RAG, KHL, YJH,SR, and SHH; Methodology: JIS, RAG, KHL, YJH, SR, and SHH; Validation: JIS, RAG, KHL, YJH,SR, and SHH; Formal Analysis: JIS, RAG, KHL, YJH,SR, and SHH; Investigation: JIS, RAG, KHL, YJH,SR, and SHH; Resources: JIS, RAG, KHL, YJH,SR, and SHH.; Data Curation: JIS, RAG, KHL, YJH,SR, and SHH; Writing – Original Draft Preparation: JIS, RAG, KHL, YJH,SR, and SHH, SY,GHJ, JLL, JYL, JWY, MAE, MIE, AK, MS, HLL, LJ, AK, JR,LS.; Writing – Review & Editing: JIS, RAG, KHL, YJH,SR, SHH, SY,GHJ, JLL, JYL, JWY, MAE, MIE, AK, MS, HLL, LJ, AK, JR,LS; Visualization, JIS, RAG, KHL, YJH,SR,SHH, SY,GHJ, JLL, JYL, JWY, MAE, MIE, AK, MS, HLL, LJ, AK, JR,LS Supervision: JIS, KHL, RAG; Project Administration JIS, KHL.

## Funding

This work was not supported by any agency or grant. No financial compensation was provided to any of these individuals.

## Acknowledgments

None

## Conflicts of interest

The authors declare no conflict of interest directly applicable to this research.

---

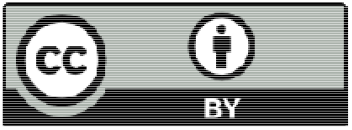

© 2020 by the authors. Submitted for possible open access publication under the terms and conditions of the Creative Commons Attribution (CC BY) license (http://creativecommons.org/licenses/by/4.0/).

## Notes

### Competing Interest Statement

The authors have declared no competing interest.

### Clinical Trial

Not an intervention study

### Funding Statement

No external funding was received

### Author Declarations

Yonsei University Medical Center, South Korea, provided an exemption because the study does not involve human or animal subjects or materials.

